# Development and internal validation of risk scores to predict survival in the pediatric population following in-hospital cardiac arrest

**DOI:** 10.64898/2026.02.01.26345326

**Authors:** Minaz Mawani, Jessica Knight, Ye Shen, Bryan McNally, Linda Brown, Mark Ebell, the American Heart Association’s Get With The Guidelines® – Resuscitation Investigators

## Abstract

**Introduction:** In-hospital cardiac arrest (IHCA) in the pediatric population is associated with poor survival and neurological outcomes. We aimed to develop and internally validate a risk score to predict survival to discharge following pediatric IHCA.

**Methods:** We included pediatric IHCA patients in the Get With The Guidelines-Resuscitation® registry between 2005 and 2021. We used logistic regression (LR), classification and regression trees (CART), and artificial neural networks (ANN) to develop models using 70% of the data and validate them using the remaining 30% of the data. Discrimination was based on the area under the receiver operating characteristic curve (AUC), and predictive accuracy on percent survival in each risk group.

**Results:** We included 6141 patients with a mean age of 4.8 years, of whom 41.3% (n = 2535) were infants < 1 year of age and 39.1% of whom survived to hospital discharge. We developed separate models for infants and older children. The most important independent pre-arrest predictors were age, illness category, acyanotic cardiac malformation, cyanotic cardiac malformation, hepatic insufficiency, hypotension/hypoperfusion, metabolic/electrolyte abnormality, metastatic/hematologic malignancy, renal insufficiency, congenital malformation, septicemia, hypotension, trauma, and pediatric cerebral performance score on admission. All three approaches showed good classification accuracy in the derivation sample (AUC for LR: 0.70, 0.71, AUC for CART: 0.68, 0.70, AUC for ANN: 0.76, 0.74 for infants and older children respectively) and used almost the same number of variables. Logistic regression and CART models were the most useful as they identified patients with the lowest survival, showed good discrimination, and could be used to develop a simple point score and decision trees that can be implemented in the clinical or research setting. In infants, the average probability of survival was 10%, 36%, and 60% whereas in older children it was 6.2%, 31.1%, and 62.3% in the low, moderate, and high survival categories in the LR model.

**Conclusion:** Pediatric patients experiencing IHCA can be classified into low, moderate, and high survival categories using a simple risk score and easily identified pre-arrest variables. These risk scores can support clinical decisions, facilitate research, and help monitor the quality of medical services.

## Introduction

In-hospital cardiac arrest (IHCA) in the pediatric population is associated with poor survival and neurological outcomes. About 2%-6% of children admitted to pediatric intensive care units are estimated to suffer from cardiac arrest and more than half of the children with IHCA do not survive to hospital discharge ^1,2,3^. About 72% of the survivors have moderate to severe functional impairment leading to diminished quality of life^4^. Identifying patients who are less likely to survive is important and could help parents with decision-making.

Risk scores, also known as prediction models or clinical prediction rules, combine multiple predictors such as patient characteristics, lab results, comorbid conditions, and other variables to estimate the probability of a clinical outcome ^5^ ^6–8^. When appropriately developed and validated, risk scores have several potential advantages over clinical judgment alone. These models can provide objective and reliable results with high accuracy while taking several factors into account as compared to clinical judgment alone which can sometimes be inconsistent and biased^9^.

Several risk scores have been developed and validated for the adult population to predict survival following IHCA, including the GO-FAR (Good Outcome Following Attempted Resuscitation), PIHCA (Prediction for outcome for In-Hospital Cardiac Arrest), and CASPRI (Cardiac Arrest Survival Post-Resuscitation In-hospital) risk scores ^10–22^. However, these were not developed for use in pediatric populations. Pediatric IHCAs are different compared to adult IHCAs in terms of etiology, comorbid conditions, severity of disease, and prognosis^2^. Risk scores that have been developed for use in pediatric populations have several methodological limitations. Holmberg and colleagues developed a risk score that predicts mortality in pediatric IHCA patients using logistic regression models. This study had a few limitations. The study included both patients with and without a loss of pulse, utilized data from 2000-2015, and included only patients with a ROSC after in-hospital arrest instead of all pulseless cardiac arrest patients ^23^. The AHA guidelines for cardiopulmonary resuscitation (CPR) and emergency cardiovascular care (ECC) were updated in 2005 with changes to CPR methods that have the potential to affect survival outcomes^24,25^. Another similar study looked at the hospital variation in survival outcomes for pediatric patients with in-hospital cardiac arrest using data from 2006 to 2010. It calculated risk-standardized survival rates at each hospital to evaluate the extent of site-level variation in outcomes. This study offers a methodological framework for future hospital comparisons of outcomes but has limited clinical applicability at the individual level^26^.

Therefore, we aimed to develop and validate a risk score to predict survival to discharge in all pediatric in-hospital pulseless cardiac arrest patients using data on pre-arrest variables from 2005 to 2021. We compared logistic regression (LR) models, classification, and regression trees (CART), and artificial neural networks (ANNs). This score has the potential to guide physicians regarding prognosis and provide a tool for evidence-based decision-making to complement their clinical judgment.

## Methods

### Study design

In this retrospective cohort study, de-identified data for model development and validation were obtained from the Get With The Guidelines® Resuscitation (GWTG-R) prospective registry from 2005 to 2021^27,28^. The Get With The Guidelines® programs are provided by The American Heart Association. This study was approved by the institutional review board (IRB) at the University of Georgia, and written informed consent was waived owing to the use of de-identified data from the registry. All participating institutions were required to comply with local regulatory and privacy guidelines and, if required, to secure institutional review board approval. Because data were used primarily at the local site for quality improvement, sites were granted a waiver of informed consent under the common rule.

### Data sources

The GWTG-R is a prospective, largest of its kind, multisite observational study of in-hospital resuscitation based on in-hospital Utstein guidelines in the US. Hospitals participating in the registry submit clinical information regarding the medical history, hospital care, and outcomes of consecutive patients hospitalized for cardiac arrest using an online, interactive case report form and Patient Management Tool^TM^ (IQVIA, Parsippany, New Jersey). IQVIA serves as the data collection (through their Patient Management Tool - PMT^TM^) and coordination center for the American Heart Association/American Stroke Association Get With The Guidelines® programs. More details can be found elsewhere ^27,28^.

### Population

Pediatric patients less than 18 years old who experienced an IHCA (absence of a palpable central pulse, unresponsiveness, and apnea) requiring either chest compressions or defibrillation or both were included in these analyses. Only the index episodes of cardiac arrest were included. Patients with missing data on survival to discharge or having a DNAR order before their index IHCA were excluded. Those episodes of arrests where the response was limited to only shock by an implantable cardioverter-defibrillator, and resuscitation at birth (e.g. delivery room arrest) were also excluded. Arrest in the inpatient settings (NICU, PICU, Pediatric CICU, Cath lab, Emergency department, general inpatient areas, Diagnostic/intervention areas, newborn nursery, OR) were included in the analysis.

### Predictor and outcome variable definitions

Standardized definitions were used for patient variables and outcomes that adhere to Utstein-style templates for cardiac arrest data^28^. Potential predictors included the following patient characteristics: age, gender, race, comorbid conditions, illness category, initial rhythm, and pediatric cerebral performance category (PCPC) score on admission. The PCPC category 1 is normal age-appropriate neurodevelopmental functioning, category 2 indicates mild cerebral disability, category 3 shows moderate disability, category 4 indicates severe disability, category 5 indicates coma/vegetative state, and category 6 indicates brain death^29^. We only included the variables for which the information is available before arrest. Definitions of the variables are provided in Supplement 1. The primary outcome for this study was a binary variable survival vs. no survival to hospital discharge. We did not include race in the model since differences in outcomes may represent racial disparities, rather than actual biological differences.

### Statistical analysis

The conduct and reporting of this study follow the transparent reporting of a multivariable prediction model for individual prognosis or diagnosis (TRIPOD) guideline^30^ (Appendix Table 1). After checking for duplicates, item analysis, and data visualization were used to assess the reliability and validity of the selected variables. New variables were defined as required by combining categories. Missing values and outliers in the data were further explored with the AHA data science team. Cases with missing information on the outcome were excluded from the analysis.

Categorical variables are presented as numbers and percentages, and continuous variables are presented as means ± SD. The significance of differences between groups with or without a risk factor for survival was determined by the Chi-square test or independent sample T-test. A two-sided P value of less than 0.05 was considered to be statistically significant. For multivariable analysis, we excluded variables exhibiting multicollinearity or with a very low prevalence. For infants, we included age, illness category, acyanotic cardiac malformation, hepatic insufficiency, hypotension/hypoperfusion, metabolic/electrolyte abnormality, respiratory insufficiency, renal insufficiency, septicemia, PCPC on admission, arrhythmia and acute non-stroke event. For older children we considered, age, illness category, acyanotic cardiac malformation, cyanotic cardiac malformation, respiratory insufficiency, acute-non-stroke, major trauma, hepatic insufficiency, hypotension/hypoperfusion, metastatic disease, metabolic/electrolyte abnormality, renal insufficiency, septicemia and PCPC on admission.

We developed separate models for infants (<1yr) and older children (1 to 17 yrs). The data was randomly divided into development (70%) and validation (30%) sets. We compared three different modeling approaches: Logistic regression (LR), classification and regression trees (CART), and artificial neural networks (ANNs). LR models make assumptions about the underlying data structure and derive adjusted odds ratio for significant risk factors which can be used to develop a simple point score, while CART models have no such assumptions and assign a probability of occurrence to each risk factor with good face validity. Therefore, CART models may outperform LR by identifying more risk factors and/or different cutoffs for abnormal within a risk factor. They also generate decision trees which are easy to understand and visualize with a tree diagram. However, they typically have lower predictive performance and can be significantly less accurate than artificial neural networks. ANNs have gained more popularity over recent years as they require little formal statistical training to develop and can learn mathematical relations between a series of input variables and corresponding output variables. These models allow more flexibility and do not require explicit distributional or other assumptions. However, they have a limited ability to identify possible causal relations, overfitting is a common problem, and they require greater computational resources. The detail of each methodology is given below.

#### Logistic regression to derive a point score-based risk prediction models

All analyses were carried out using SAS 9.4 (SAS Institute Inc, Cary, North Carolina, USA). We used the backward elimination method to identify the most suitable predictors of survival to build the model. A backward selection model considers the effects of all the variables simultaneously and sequentially removes them until the AIC or BIC cannot further improve. A P-value <0.05 was considered statistically significant. For the ease of score development, we converted continuous into categorical variables. We report adjusted odds ratios with 95% confidence intervals. The rounded beta coefficients from the final model were used to construct the score. We developed point scores by dividing the beta coefficients by the smallest beta coefficient. The scores were divided into low, moderate, and high likelihood of survival categories^31,32^. Discrimination was evaluated using AUROC whereas, calibration was evaluated using Hosmer-Lemeshow statistic and calibration plots of observed vs. expected survival.

#### Classification and Regression Trees (CART) based prediction models

All CART analyses were carried out using JMP Pro 17.2.0 (SAS Institute Inc, Cary, North Carolina, USA). Classification and regression tree (CART) analysis was used to develop a model predicting survival outcomes. The algorithm uses non-parametric tests and recursive partitioning to evaluate data and progressively classify patients into subgroups based on the optimal independent predictors. The best predictor variable is determined by calculating the likelihood ratio chi-square test for each possible split. The variables and discriminatory values used and the order in which the splitting occurs are produced by the underlying mathematical algorithm and are calculated to maximize the resulting predictive accuracy. The algorithm identifies the predictor variable that best discriminates between patients with and without the outcome. We considered all important variables for the CART models. To optimize the performance of the model for future data and avoid overfitting, a combination of stopping rules and pruning were applied to the tree. We restricted that a minimum of 50 observations be allowed in any terminal node. The unsupervised modeling approach automatically selected the splitting variable based on the log worth statistic and continued until there was no improvement in the AUROCC (area under the receiver operating characteristic curve). Ends with similar probabilities of the outcome were grouped to form patient groups with low, moderate, and high probability of survival to discharge.

#### Supervised machine learning models using artificial neural networks (ANN)

We carried out analyses using RStudio (version 2023.06.0; The R Foundation, Vienna, Austria). We used supervised machine learning and ANNs to predict survival using a classification neural network model using ‘neural net’ package in R. Feature selection was performed using correlations. The network consisted of an input layer, hidden layers, and an output layer. The input layer consisted of all pre-arrest variables. Hidden layers with different numbers of nodes were tested. The output layer contained only one node for classifying the binary outcome of survival to discharge. We used a logistic activation function. For the output layer, the sigmoid activation function was used because of the binary outcome variable. Model tuning was performed to find an optimal model by tuning different hyperparameters to find a model with the least error. The model was tested on validation data and patients were classified into low moderate and high survival categories.

### Model performance

Model performance for all three methods (LR, CART, and ANN) was evaluated using discrimination (area under the receiver operating characteristic curve or AUROCC) and the ability of the model to classify patients in low, moderate, and high survival groups. Results were compared to find the simplest model with optimum performance. All statistical tests were two-sided, we considered P <0.05 to be statistically significant.

## Results

### Study population

Our data included 6141 pediatric patients who experienced IHCA (Strobe flowchart S1 in Supplement). Our study sample represented data from 49 states, mostly urban (99%), and mostly from large (58.6%) or medium (41.2%) sized JCAHO certified hospitals. Most of the hospitals were teaching hospitals (99%) with an average daily census of 401 patients (range: 21 – 991). The characteristics of all included patients by survival are presented in Table 1. Our study population was primarily male (54.7%) with an average age of 4.8 years. Most patients were white (41.7%) followed by black (23.2%), Hispanic (18.3%), and others (17.4%). Most of the arrests (87.1%) occurred in a monitored location and 2398 (39.1%) patients survived to hospital discharge, of those 1114 (46.3%) had a normal neurological status or mild cerebral disability (PCPC 1 or 2).

**Table 1:**
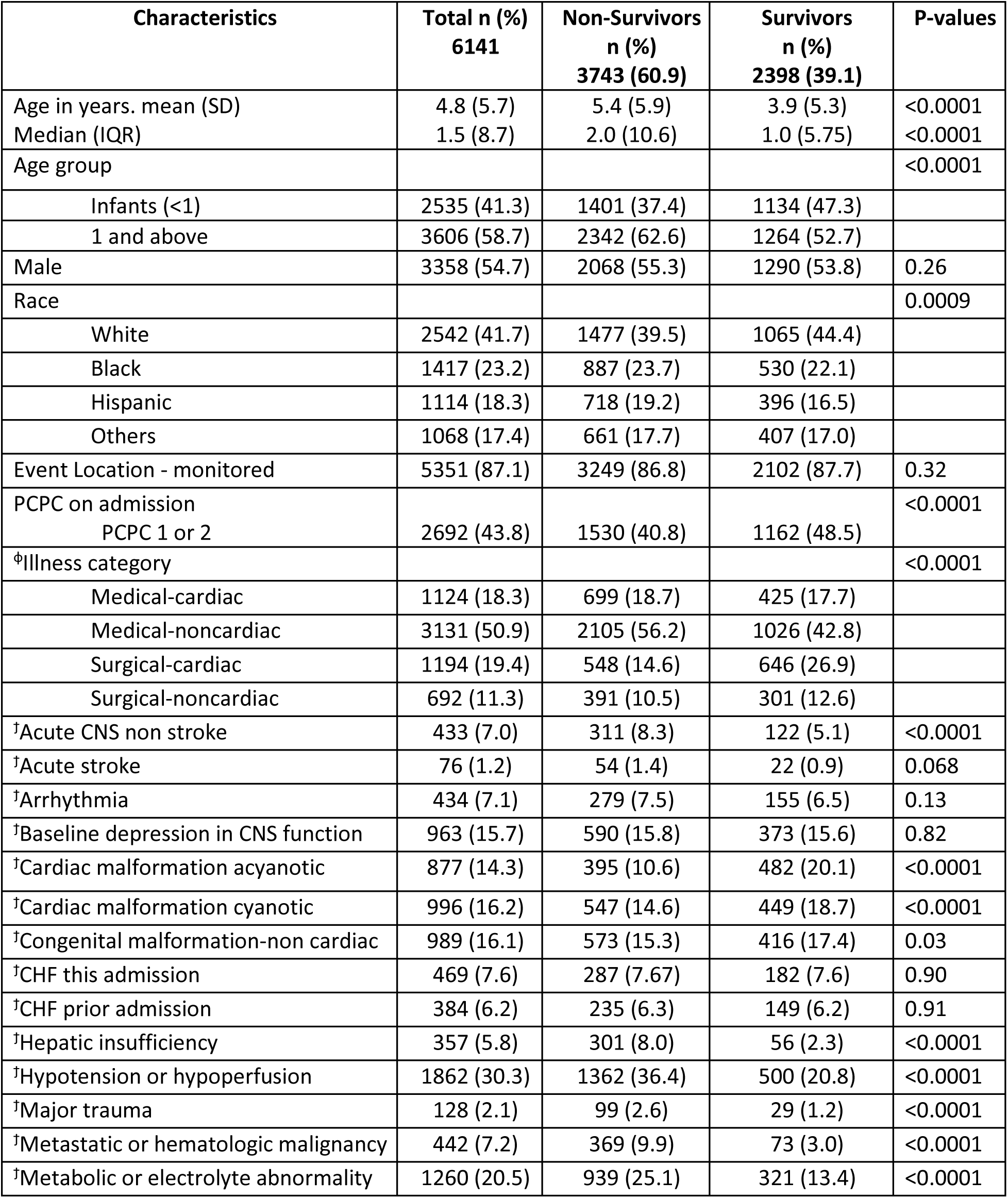
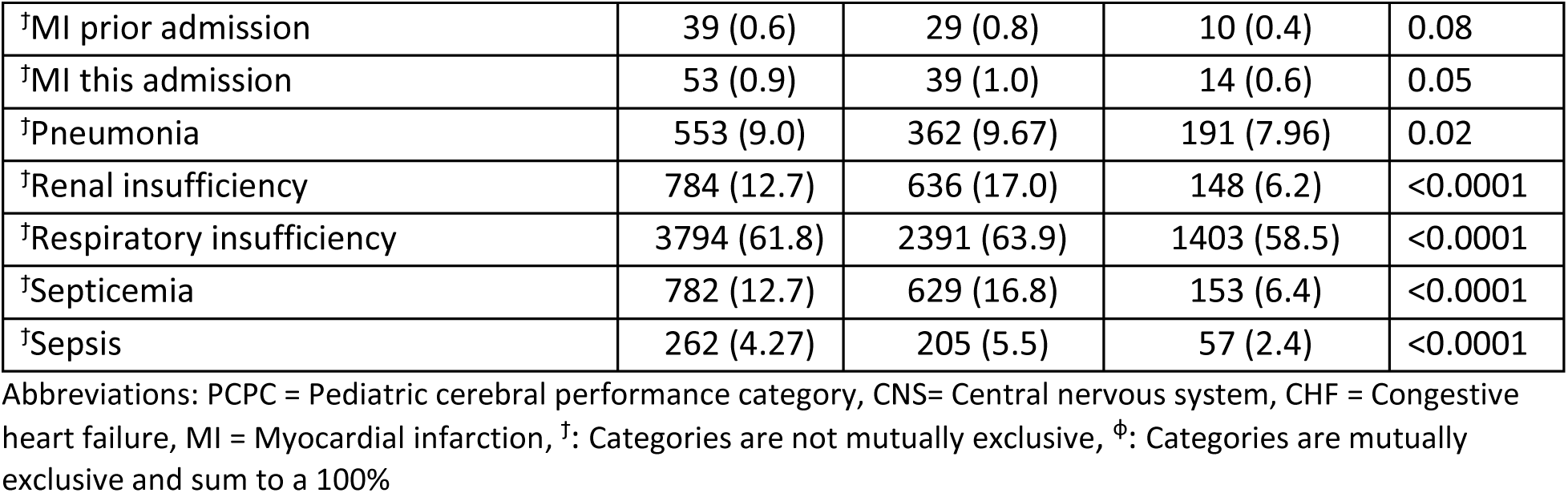
Comparison of characteristics between survivors vs. non-survivors for all children (n = 6141)

Survivors were younger than non-survivors with a mean age (SD) of 3.9(5.3) vs. 5.4(5.9). Survival was the highest for White (41.9%) followed by Black (37.4%) and Hispanic (35.5%) patients. Survival was highest for patients with a primary admitting diagnosis of surgical cardiac illness (54.1%). We developed separate models for infants and older children because of differences in characteristics, comorbid conditions, and survival outcomes across these two populations (Table S1).

### Logistic regression models

Tables 2 and 3 describe the adjusted odds ratios with 95% confidence intervals for the logistic regression models for infants and older children respectively.

**Table 2.** Factors associated with survival to discharge using logistic regression model in infants using development sample (n = 1775)

| Characteristics | B-coefficients | Point scores | Adjusted Odds ratios (95%CI) |
| --- | --- | --- | --- |
| Age category |  |  |  |
| Neonates (< 1 month) | - |  | Ref |
| Infants (1 month to <1 year) | 0.38 | 1 | 1.47 (1.17-1.84) |
| Illness category |  |  |  |
| Medical-cardiac | 0.004 | 0 | 1.01 (0.67-1.51) |
| Medical-noncardiac | 0.29 | 1 | 1.34 (0.93-1.94) |
| Surgical-cardiac | 0.48 | 2 | 1.62 (1.10-2.39) |
| Surgical-noncardiac | Ref | 0 | Ref |
| Cardiac malformation- acyanotic | 0.61 | 2 | 1.85 (1.42-2.40) |
| Hepatic insufficiency | -0.85 | -3 | 0.43 (0.23-0.80) |
| Hypotension/hypoperfusion | -0.69 | -2 | 0.50 (0.39-0.64) |
| Metabolic electrolyte abnormality | -0.49 | -2 | 0.61 (0.46-0.82) |
| Renal insufficiency | -0.73 | -3 | 0.48 (0.32-0.73) |
| Septicemia | -1.07 | -4 | 0.34 (0.23-0.51) |
| Admission PCPC 1 or 2 | 0.43 | 1 | 1.54 (1.26-1.90) |

Abbreviations: CI = Confidence intervals, PCPC = Pediatric cerebral performance category, Ref = Reference category, OR here refers to odds of survival.

**Table 3.** Factors associated with survival to discharge using logistic regression models in older children using development sample (n = 2525)

| Characteristics | B-coefficients | Point score | Adjusted Odds Ratios (95% CI) |
| --- | --- | --- | --- |
| Age category |  |  |  |
| Children (1 to < 12 years) | 0.37 | 2 | 1.44 (1.20-1.76) |
| Adolescents (12 to <18 years) | Ref |  | Ref |
| Illness category |  |  |  |
| Medical-cardiac | -0.76 | -4 | 0.47 (0.34-0.65) |
| Medical-noncardiac | -0.98 | -5 | 0.37 (0.28-0.49) |
| Surgical-cardiac | 0.18 | 1 | 1.20 (0.81-1.77) |
| Surgical-noncardiac | Ref | 0 | Ref |
| Cardiac malformation acyanotic | 0.30 | 2 | 1.35 (1.001-1.83) |
| Cardiac malformation cyanotic | -0.41 | -2 | 0.66 (0.47-0.92) |
| Hepatic insufficiency | -1.04 | -6 | 0.35 (0.21-0.59) |
| Hypotension/hypoperfusion | -0.70 | -4 | 0.49 (0.40-0.61) |
| Major trauma | -0.84 | -5 | 0.43 (0.23-0.79) |
| Metastatic malignancy | -0.86 | -5 | 0.42 (0.29-0.60) |
| Renal insufficiency | -0.50 | -3 | 0.61 (0.45-0.82) |
| Septicemia | -0.54 | -3 | 0.58 (0.43-0.79) |
| Admission PCPC 1 or 2 | 0.22 | 1 | 1.24 (1.04-1.49) |

Abbreviations: CI = Confidence intervals, PCPC = Pediatric cerebral performance category, Ref = Reference category

#### Infants

The use of backward regression yielded a model comprising of 9 pre-arrest predictor variables in infant population (Table 2). Being older than a month, having a surgical-cardiac or surgical non-cardiac primary diagnosis, acyanotic cardiac malformation, or having a PCPC 1 or 2 at admission were associated with survival. Comorbid conditions such as hepatic insufficiency, hypotension or hypoperfusion, metabolic or electrolyte abnormality, renal insufficiency, or septicemia pre-arrest were associated with non-survival. The resulting model had an AUROCC of 0.70. We developed point scores by dividing the beta coefficients by the second smallest beta coefficient (0.29) to develop the score in this population since using the smallest beta resulted in very widely spread-out scores. The development sample included 1775 infants, with 796 (44.8%) surviving to discharge. The validation sample included 760 infants, with 338 (44%) surviving to discharge. The performance of the risk score in development and validation samples is described in Table 4. The model classified about 11.5% of patients in the low survival group, 40% in the moderate survival group, and 48% in the high survival group with an average probability of survival of 10%, 36%, and 60% in the low, moderate, and high survival categories. The results were similar in the validation group. Evaluation in the validation group yielded an AUROCC of 0.66 The model had good calibration as evidenced by calibration plots with an insignificant p-value for Hosmer and Lemeshow. (Figure S2).

**Table 4:** Classification accuracy of the risk prediction models in the development and validation groups using logistic regression.

|  | Development group |  | Validation group |  |
| --- | --- | --- | --- | --- |
| Risk Category (Points) | Survival n/Total (%) | LR | Survival n/Total (%) | LR |
| **Infants** |  |  |  |  |
| Low survival (-14 to -4) | 21/204 (10.2) | 0.14 | 8/76 (10.5) | 0.14 |
| Moderate survival (-3 to 1) | 256/711 (36.0) | 0.69 | 121/320 (37.8) | 0.75 |
| High survival (2 to 6) | 519/860 (60.3) | 1.87 | 209/364 (57.4) | 1.68 |
| Total | 796/1775 (44.8) |  | 338/760 (44.5) |  |
| **Older children** |  |  |  |  |
| Low survival (-25 to -13) | 12/194 (6.2) | 0.13 | 9/83 (10.8) | 0.21 |
| Moderate survival (-12 to -1) | 597/1917 (31.1) | 0.86 | 283/813 (34.8) | 0.92 |
| High survival (0 to 6) | 258/414 (62.3) | 3.16 | 105/185 (56.8) | 2.26 |
| Total | 867/2525 (34.3) |  | 397/1081 (36.7) |  |

Abbreviations: LR = Likelihood ratios

#### Older children

In older children, the backward regression yielded a model comprising 11 pre-arrest predictor variables (Table 3). Age < 12 years, having a primary diagnosis of surgical cardiac, acyanotic cardiac malformation and PCPC 1 or 2 on admission were associated with survival, whereas comorbid conditions such as cyanotic cardiac malformation, hepatic insufficiency, hypotension, major trauma, metastatic malignancy, renal insufficiency, septicemia, medical primary diagnoses were associated with non-survival. This model had an AUROCC of 0.71. The development sample included 2525 patients, with 867 (34.3%) surviving to discharge, whereas the validation sample included 1081 patients, with 397 (36.7%) surviving to discharge. The performance of this risk score is described in Table 4. The model classified 7.7%, 76.0%, and 16.4% in low, moderate, and high survival categories with approximately 6.2%, 31.1%, and 62.3% average probability of survival in each group respectively. Evaluation in the validation group yielded an AUROCC of 0.67 with a good model calibration(Figure S2).

### CART models

#### Infants

The model development sample consisted of 1775 patients with 790 (45%) surviving to hospital discharge and yielded an AUROCC of 0.68 using 14 splits and 15 terminal nodes (Figure 1). The model used 10 pre-arrest variables such as hypotension, metabolic electrolyte abnormality, septicemia, renal insufficiency, illness category, PCPC on admission, age in years, respiratory insufficiency, acyanotic cardiac malformation, and congenital malformation to classify patients in low, moderate, and high survival categories. Hypotension was considered the most important variable by the CART algorithm. The algorithm chose age cutoffs of 6 days and 5 months at separate instances as optimal cut-offs for the split. Compared with the logistic regression model, congenital malformation and respiratory insufficiency were added as important predictor variables whereas hepatic insufficiency was dropped by the CART algorithm. We manually pruned the model to remove nodes that did not have clinically meaningful differences in survival between paired terminal nodes. We combined terminal nodes to create categories for low (11% to 16%), moderate (24% to 30%), and high (42% to 69%) likelihood of survival. The model classified 11.7%, 15.3%, and 72.9% of patients in the low, moderate, and high survival categories with an average probability of survival of 13.3%, 26.4%, and 53.3% in each category respectively (Table 5 and Table S2). Evaluating the CART model in the validation sample yielded an AUROCC of 0.68 (Figure S2).

**Figure 1:**
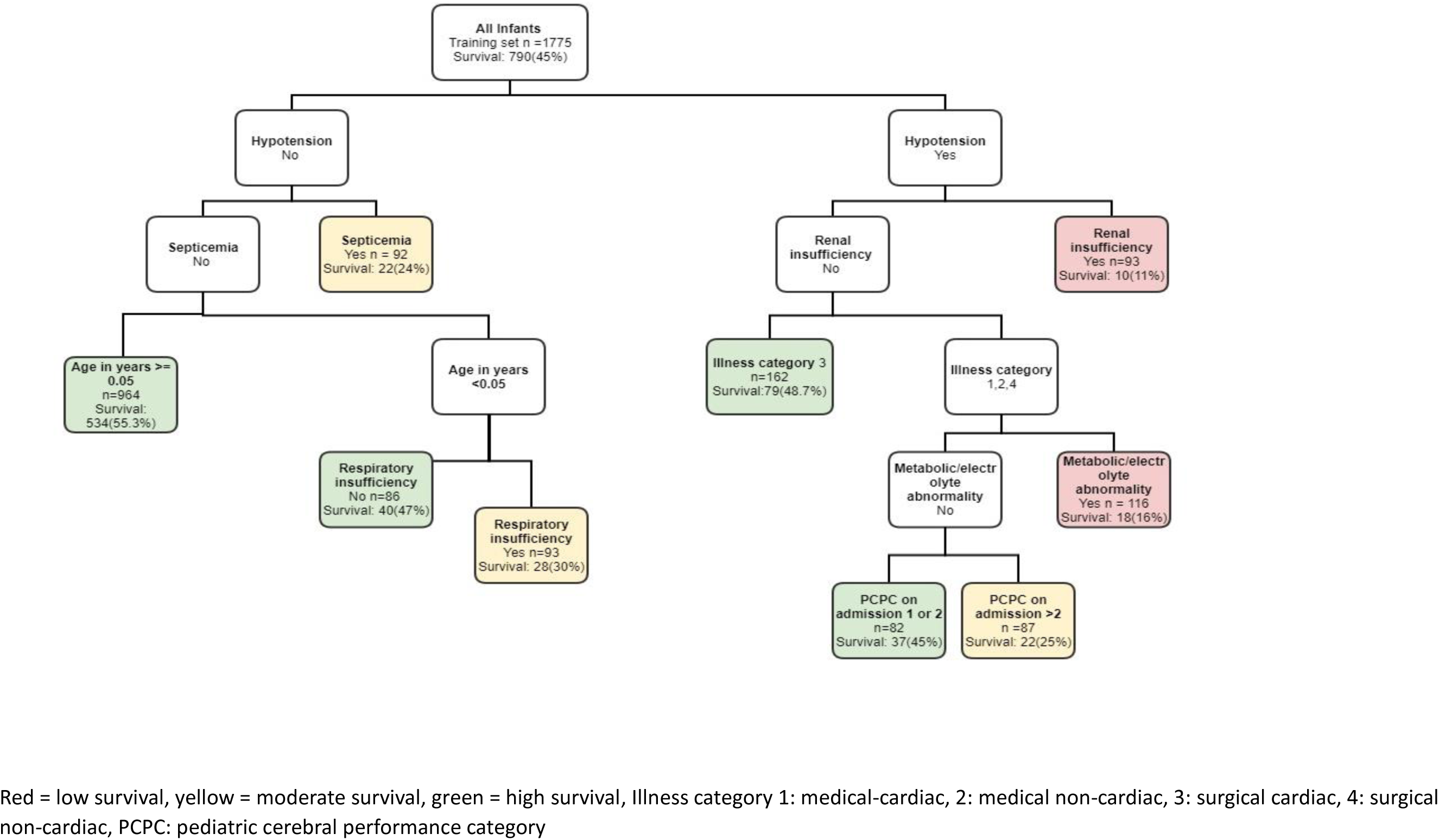
Unsupervised CART models for in-hospital cardiac arrest, estimating survival to discharge in infants.

**Table 5.** Overall comparison of logistic regression, classification and regression trees and artificial neural networks.

|  | Logistic Regression |  | Classification and Regression Trees |  | Artificial Neural Networks |  |
| --- | --- | --- | --- | --- | --- | --- |
|  | Deaths/Total (%) |  | Deaths/Total (%) |  | Deaths/Total (%) |  |
|  | Infants | Older children | Infants | Older children | Infants | Older children |
| AUROC |  |  |  |  |  |  |
| Derivation | 0.70 | 0.71 | 0.68 | 0.70 | 0.76 | 0.74 |
| Validation | 0.66 | 0.67 | 0.68 | 0.68 | 0.66 | 0.67 |
| Number of Predictors | 9 | 11 | 10 | 9 | 9 | 9 |
| Survival % by risk group -Derivation |  |  |  |  |  |  |
| Low survival | 21/204 (10%) | 12/194 (6.2%) | 28/209 (13.3%) | 42/346 (12%) | 63/443 (14.2%) | 79/632 (12.5%) |
| Moderate survival | 256/711 (36%) | 597/1917 (31.1%) | 72/272 (26.4%) | 265/1042 (25%) | 244/620 (39.3%) | 390/1261 (30.9%) |
| High survival | 519/860 (60%) | 258/414 (62.3%) | 690/1294 (53.3%) | 564/1136 (49.6%) | 488/710 (68.7%) | 386/630 (61.2%) |
| Survival % by risk group - Validation |  |  |  |  |  |  |
| Low survival | 8/76 (11%) | 9/83 (10.8%) | 15/100 (15%) | 24/151 (16%) | 43/190 (22.6%) | 57/271 (26.6%) |
| Moderate survival | 121/320 (38%) | 283/813 (34.8%) | 36/131 (27.4%) | 117/439 (27%) | 114/266 (42.8%) | 194/539 (35.9%) |
| High survival | 209/364 (57%) | 105/185 (56.8%) | 293/529 (55.3%) | 252/492 (51.2%) | 182/304 (59.6%) | 158/271 (58.3%) |
| % of patients in each risk group - derivation |  |  |  |  |  |  |
| Low survival | 11.5% | 7.7% | 11.7% | 13.7% | 25% | 25% |
| Moderate survival | 40.1% | 76% | 15.3% | 41.2% | 35% | 50% |
| High survival | 48.5% | 16.4% | 72.9% | 45% | 40% | 25% |

Abbreviations: AUROC = Area under the receiver operating characteristic curve

#### Older children

The model for the development sample comprised 2524 patients with 871 (35%) surviving to hospital discharge. It used 18 splits and 19 terminal nodes and yielded an AUROCC of 0.70 (Figure 2). The model used 9 pre-arrest variables including Illness category, renal insufficiency, metastatic malignancy, hypotension, congenital malformation, respiratory insufficiency, PCPC on admission, age in years, and metabolic electrolyte abnormality to classify patients in survival categories. The illness category was considered the most important variable by the algorithm. The algorithm selected the age cut-off of 1.08 years. Compared with logistic regression, the CART models added respiratory insufficiency, and congenital malformation, and did not include major trauma, cyanotic cardiac malformation, acyanotic cardiac malformation, septicemia, and hepatic insufficiency. We combined terminal nodes to create categories of low (4% to 15%), moderate (18% to 33%), and high (41% to 73%) survival categories. The model classified 13.7%, 41.2%, and 45% of patients in the low, moderate, and high survival categories and had an average probability of survival of 12%, 25%, and 49.6% in each group respectively (Table 5 and Table S2). Evaluating the CART model in the validation sample yielded an AUROCC of 0.68 (Figure S2).

**Figure 2:**
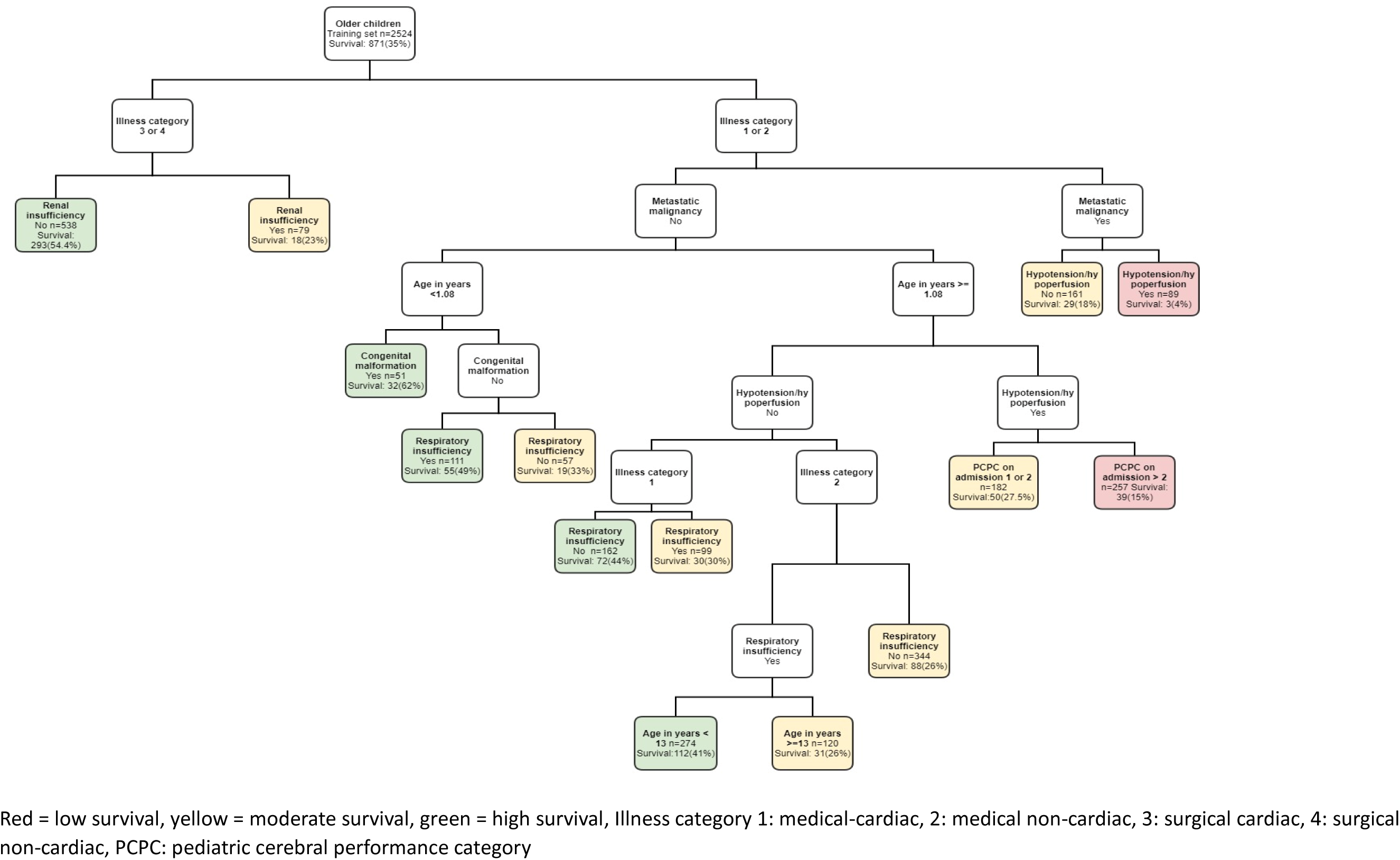
Unsupervised CART models for in-hospital cardiac arrest, estimating survival to discharge in older children.

### Artificial neural networks

#### Infants

The model derivation group consisted of 1774 patients. Feature selection identified 9 pre-arrest variables such as age, illness category, acyanotic cardiac malformation, hepatic insufficiency, hypotension, metabolic abnormality, renal insufficiency, septicemia, and PCPC on admission. We used resilient backpropagation, logistic activation function, and a learning rate of 0.1. Different iterations were performed to select a model with optimum performance and the least error in the derivation population. Our final model had two hidden layers with 5 and 3 nodes. The patients were classified into low, moderate, and high survival categories and the model was tested on a validation sample (n=761). The AUROCC for model development and validation was 0.76 and 0.66 respectively. The model classified 25%, 35%, and 40% of patients in the low, moderate, and high survival categories with an average probability of survival of 14.2%, 39.3%, and 68.7% within each category respectively (Table 5 and Table S3).

#### Older children

There were 2524 and 1082 patients in the development and validation samples. Feature selection identified 9 pre-arrest variables: age, illness category, acyanotic cardiac malformation, hepatic insufficiency hypotension, metastatic malignancy, renal insufficiency, septicemia, and PCPC on admission. We used resilient backpropagation, logistic activation function, and a learning rate of 0.1. Different iterations were performed to select a model with optimum performance and the least error. Our final model had two hidden layers with 5 and 3 nodes, a learning rate of 0.1, and an AUROCC of 0.74. The patients were classified into the low, moderate, and high survival categories and the model was tested on a validation sample (n=1082). The model classified 25%, 50%, and 25% of the patients in low, moderate, and high survival categories with an average survival probability of 12.5%, 30.9%, and 61.2% within each category respectively (Table 5 and Table S3). The AUROCC for the validation model was 0.67 (Figure.S1). There was a large difference in survival for the low category across the development and validation sample (>8%) in both the models for infants and older children.

## Discussion

We developed and internally validated simple risk scores to predict survival to discharge in pediatric in-hospital cardiac arrest patients using three different statistical approaches, logistic regression, classification and regression trees, and artificial neural networks. We developed separate models for infants and older children using the three methods that classified children into categories of low, moderate, and high likelihood of survival.

We found logistic regression and CART models to be most useful. CART models are simpler to follow because of the flow-chart presentation and classified more patients in the low and high survival categories while using the same number of predictors, they also had a comparable accuracy to LR models. Logistic regression models identified patients with the lowest survival, had good discrimination for the development sample, and had close enough results in the validation sample indicating a higher possibility of good performance on unseen data. It also provided odds ratio while adjusting for all other variables, provided a more intuitive explanation, and could be used to develop a simple point score that can be implemented in research settings. In addition, logistic regression models have a low variance, and the model predictions are relatively stable when applied to different subsets of the same dataset or used on entirely new data set.^9,33–35^

The artificial neural network was useful in classifying more patients in the low survival categories, compared to LR and CART, and had the highest accuracy in the development sample compared to the other two models. However, the performance deteriorated in the validation sample indicating overfitting and there was a much higher rate of survival in the low-risk group in the validation sample which is less clinically helpful. ANN models are prone to overfitting as it causes the model to memorize the noise and reduce its accuracy, which may explain the loss of accuracy with validation. The ANNs also represent a black box phenomenon – they provide limited insight into a problem, have a limited ability to explicitly identify possible causal relations, are hard to use at the bedside, and require greater computational resources.

These risk scores can provide quick and inexpensive estimates of the probability of survival ^6^. These can be of great value in assisting clinical decision-making and can complement clinical opinion and intuition. The awareness and understanding of what predictive factors play an important role in achieving a favorable outcome are relevant for the acceptance of poor outcomes following IHCA by the relatives ^36^. The purpose of developing these clinical prediction rules was to identify groups with very low survival to help guide DNAR decisions. We could not do that because of an overall high survival for this registry. However, this tool can be a valuable resource in several ways besides prognostication. It can help stratify patients for clinical trials by identifying patients in particular risk categories who may be more or less likely to benefit from a specific intervention. It can help facilitate research across institutions by improving the homogeneity of patient groups and can help develop evidence-based guidelines. It can also help monitor the quality of medical services^6–8^.

Predictors of survival identified in our study were largely consistent with previous similar studies^23^. We found higher survival rates for infants compared with older children^37^. We also report a better survival for surgical cardiac patients compared to other groups^38^. Our study found acyanotic cardiac malformation to be associated with survival. While this may seem counterintuitive, it may be because these patients are more likely to be admitted to a monitored unit.

This study has several strengths. To the best of our knowledge, this is the first study developing a risk score to predict the outcomes of pediatric in-hospital cardiac arrest patients using easily available pre-arrest variables from a nationwide population-based registry. Our study sample was largely representative of the country. We developed separate models for infants and older adults because of inherent differences in characteristics and outcomes for these populations. We used separate samples to develop and internally validate the score which ensures the accuracy and stability of the models.

This study has a few limitations. A meaningful survival outcome for a prediction model would be whether the surviving child is neurologically functional and will have a good quality of life. However, it was not the scope of the present study We adjusted for PCPC on admission, however, this scale has been found be inadequate in capturing neurological impairments in critically ill neonates and infants^39^. We did not adjust for gestational age in our analysis. Our AUROCC indicated fair performance which could be because of unmeasured confounding. Besides the pre-arrest variables that we used, other factors such as pre-arrest labs, sociodemographic, and other factors could be important predictors and were not measured by the registry.

The definitions used for low, moderate, and high survival in our study were arbitrary since no previous work has identified cut-offs for futility with reference to pediatric survival, and cut-offs from the adult population could not be used. More work is needed to identify medical futility for this population based on the preferences of physicians and parents. There is a potential for selection bias for this dataset since those with poor survival and DNAR status were already excluded from the registry. This could be the reason we did not find a very low survival population in our data. In addition, the registry represents data from mostly tertiary care teaching hospitals which also limits the generalizability of these findings. Our models measured survival to discharge, and we do not know how many of these would survive long-term, 3, or 6 months post-discharge, and it would be relevant to measure that in future studies. Finally, there is a need to prospectively validate this model in the same population using the data from 2022 to 2025 to account for changing survival. In addition, it would be important to validate this model in a different population with different prevalence. For instance, testing it in data from hospitals that do not participate in the AHA GWTG-R registry.

## Conclusion

We have successfully developed and internally validated a risk prediction score that is easy to use, transparent, and with good accuracy. Our model could not identify patients with a very low probability of survival. This tool will need to be externally validated before implementing in a clinical setting. This tool could be helpful in strafing patients for research and quality improvement projects besides prognostication.

## Data Availability

Data will be available upon a reasonable request

## Acknowledgements

American Heart Association’s Get With The Guidelines®-Resuscitation Pediatric Research Task Force members: Anne-Marie Guerguerian MD PhD FRCPC FAAP FAHA; Caitlin E. O’Brien MD MPH; Ericka L. Fink MD MS; Javier J. Lasa MD FAAP; Joan S. Roberts MD; Lillian Su MD; Linda L. Brown MD MSCE; Maya Dewan MD MPH; Monica Kleinman MD; Noorjahan Ali MD MS FAAP; Punkaj Gupta MBBS; Robert M. Sutton MD MSCE; Ron Reeder MS PhD; Todd Sweberg MD MBA

## Data Sharing

Study data, protocol and analysis code will be available on a reasonable request. The study was not required to be registered.

## Patient and Public Involvement

Patient/public was not involved in the design, conduct, reporting, interpretation or dissemination of the study

